# Quantitative risk assessment of COVID-19 and serious illness among spectators at mass gathering events with vaccine-testing package implementation

**DOI:** 10.1101/2022.01.30.22269980

**Authors:** Michio Murakami, Tsukasa Fujita, Yuichi Iwasaki, Masaki Onishi, Wataru Naito, Seiya Imoto, Tetsuo Yasutaka

## Abstract

While mass gathering events have resumed in conjunction with vaccine-testing (VT) packages, their effects on reducing COVID-19 risk remain unclear. Here, we used an environmental exposure model to analyze the effects of vaccinations and proof of negative test results on reducing infection risk and serious illness among spectators at mass gathering events. We then analyzed the difference in risk with and without VT and regular seat zoning. Risk of infection and serious illness were quantified using a model incorporating parameters such as vaccination coverage, vaccine prevention effectiveness, and sensitivity of polymerase chain reaction (PCR) or qualitative antigen tests. When vaccine prevention effectiveness was 50% (corresponding to 4 months for the delta variant and 1–2 months for the omicron variant after the second vaccine dose), the risk of infection and serious illness among vaccinated spectators were 0.32–0.40 and 0.13–0.16 times of those who tested negative, respectively. In contrast, the risks of infection and serious illness among vaccinated spectators without measures such as mask wearing were 4.0 and 1.6 times higher than those among unvaccinated spectators with such measures, respectively. The risk of infection with an 80% vaccination coverage and a vaccine prevention effectiveness of 20% (corresponding to 5–6 months for the delta variant or 3–4 months for the omicron variant after the second vaccine dose) was comparable to that of a 20% vaccine coverage and a vaccine prevention effectiveness of 80% (corresponding to 1–3 months for delta variant after the second vaccine dose). Regarding zoning, there was little difference in risk with a vaccination coverage of ≥80%. Adherence to individual measures after vaccination and maintenance of high vaccine effectiveness among spectators at stadiums are important for reducing risk of infection and serious illness. Furthermore, seat zoning did not affect overall infection risk reduction.

## 1. Introduction

Since the coronavirus disease 2019 (COVID-19) global pandemic started, balancing infection control and social activities in daily life has become an important issue. Mass gatherings, such as sports events, were initially considered one of the modes through which transmission of COVID-19 occurred (Stange et al., 2021), and were therefore required to be cancelled or postponed in the first half of 2020 (Parnell et al., 2020). Subsequently, depending on infection levels, mass gathering events were gradually initiated again with various measures at stadiums in place, such as mask wearing, ventilation, disinfection, and capacity limitations (Chiampas and Ibiebele, 2021). In a randomized controlled trial, empirical case surveys observed COVID-19 risk of infection among spectators at a mass gathering event (Revollo et al., 2021). Another study investigated the differences in the risk of infection with and without measures, including mask wearing (The United Kingdom Government, 2021a). A numerical simulation-based study using an environmental exposure model estimated that a combination of various measures, such as mask wearing and ventilation, can achieve a 99% risk reduction among spectators participating in mass gathering events (Murakami et al., 2021a). The proportion of adherence to these measures has also been measured in actual events (Murakami et al., 2021b).

With increased COVID-19 vaccination coverage (VC), behavioral restrictions have been lifted on vaccinated individuals in various countries, such as the United States and United Kingdom (Sleat et al., 2021). In addition, due to ethical considerations regarding unvaccinated individuals, some countries have utilized requiring proof of negative test results in place of vaccination proof, referred to as vaccine-testing (VT) package. Similarly, Germany lifted restrictions on social activities, including mass gathering events, for people who are either vaccinated, have been infected, or have a negative test result (known as the “3G rule”) (Desson et al., 2021). This was subsequently changed to the “2G rule,” excluding those with a negative test result (BBC, 2021). On November 16, 2021, the Subcommittee on Novel Coronavirus Disease Control in Japan proposed that even in worsening infection situations, such as under a declared state of emergency, the VT package system should be applied to allow spectators to attend events to stadium capacity, as long as a safety plan for preventing infections had been established, such as droplet control measures (Subcommittee on Novel Coronavirus Disease Control, 2021). The Cabinet Office announced the implementation of the VT package proposal on November 19, 2021(Cabinet Secretariat, 2021), however, on January 19, 2022, it was paused in principle due to the omicron variant emergency (Advisory Committee on the Basic Action Policy, 2022).

The effectiveness of the VT package in reducing the risk of infection remains unclear. In the United Kingdom, where vaccination efforts have progressed, more than 3,000 cases of infection occurred during the UEFA Euro 2020 Final that took place on July 11, 2021, in which many participants did not take measures such as mask wearing (Smith et al., 2021). Previous studies show that anti-spike immunoglobulin G and neutralizing antibody levels decline even after the second dose of the BNT162b2 vaccine (Pfizer-BioNTech) (Levin et al., 2021). Furthermore, with the emergence of the B.1.351 (beta) and B.1.617.2 (delta) variants, vaccine prevention effectiveness (VPE) against infection is reduced 4 months after the second dose, while VPE against serious illness is maintained for more than 6 months (Chemaitelly et al., 2021). It is necessary to evaluate how much the risk of infection and serious illness can be reduced by vaccination and testing for negative results, and how effective these are compared to other measures at stadiums, such as mask wearing. Furthermore, VT package implementation poses challenges in the practical operations of organizing mass gathering events. During mass gathering events in Japan, VT packages are not necessarily used for all seats, but have been partly applied to some seats with the reservation request of the users. However, the extent to which the risk of infection differs with and without the zoning of VT package seats (i.e., seats for users of VT packages) and regular seats (i.e., seats for non-users of VT packages) remains unclear.

The purpose of this study was three-fold. First, we estimated the effects of vaccination and proof of negative test results on reducing the risk of infection and serious illness among spectators at mass gathering events with and without various measures in place at stadiums. Second, we assessed the extent to which the risk of infection among spectators differed with variations in VC and VPE against infection. Third, we analyzed how the risk of infection and serious illness differed with and without zoning of VT and regular seats. This study aimed to perform a quantitative risk assessment on how mass gathering events should be conducted during the COVID-19 epidemic while considering practical components on the use of VT packages.

## 2. Method

### 2.1. Model

In this study, we used a modified environmental exposure model (Murakami et al., 2021a; Yasutaka et al., 2021). In the model, the amount of exposure through four routes (direct droplet spray, direct inhalation of inspirable particles, hand contact, and inhalation of respirable particles) and the corresponding risk of infection were estimated after considering factors such as the amount of virus emitted by infectors, changes in virus concentrations in the environment, and surface transfer. Furthermore, it evaluated and quantitatively assessed various measures for the reduction in the amount of virus at stadiums, such as mask wearing, ventilation, and disinfection. The proportion of asymptomatic infectors of COVID-19 among spectators entering the stadium was considered an experimental condition in the previously mentioned model. The environmental dynamics of the virus were calculated for the stadium, which was divided into two areas: the stands and other locations (concourse, restrooms, and concession areas). The parameters for estimating environmental virus concentration and exposure included the frequency and saliva volume of talking, coughing, and sneezing; virus concentration in saliva; viral viability ratio; air exchange rate; inactivation rate; surface contact frequency; and transfer rate by contact. The parameters of the dose-response relationships were based on a previous study (k = 410) that examined the number of different doses in mice and the risk of severe acute respiratory syndrome coronavirus (SARS-CoV) infection (Watanabe et al., 2010), which was similar to the parameter value for SARS-CoV-2 (k = 530 at airborne particle contribution levels of 0.5) obtained from limited experiments on ferrets as well as estimates of human exposure (Zhang and Wang, 2021).

Measures included wearing unwoven masks to reduce virus emission and frequency of contact with the face, ventilation to reduce airborne virus concentrations, physical distancing at entry and exit, disinfection to inactivate viruses on environmental surfaces, hand washing to inactivate and remove viruses from fingers, and wearing headwear to protect against contact with hair. In the scenario with measures in place, it was assumed that all measures were taken together.

There were five categories of spectators: (1) individuals who accompanied the infector, (2) persons sitting immediately in front of the infector in the stands, (3) persons exposed in restrooms after use by the infector, (4) persons exposed in concession areas after use by the infector, and (5) others.

The modifications from the previous model (Murakami et al., 2021a) have been described in a previous report (Yasutaka et al., 2021) and are as follows: The time spent in the stands was set to 3 h (1 h before the game and 2 h during the game) and the time spent in the concourse (for entry), restrooms, concession areas, and concourse (for exit) was set at 15 min each, for a total time of 4 h (football conditions). The number of spectators and proportion of the capacity used were set at 40,000 people and 100%, respectively. The number of persons accompanying the infector was set at 50% probability of being 2, 35% probability of being 1, and 15% probability of being 0. The probability of the infector talking in the stands per minute was set at 0.3 before the game and 0.2 during the game without measures, while 0.15 and 0.1 were the probabilities set for before and during the game with measures, respectively, under the assumption that the spectators refrained from talking. The probabilities of the infector facing the front, left, and right sides of the stands were set to 0.7, 0.15, and 0.15, respectively. Instead of setting up partitions in the stands, it was assumed that masks would be worn even in the stands.

In addition, we made the following changes from our previous study (Yasutaka et al., 2021). First, the proportion of asymptomatic infectors among spectators entering the stadium was considered an experimental parameter in the previous study, while the daily incidence of new infections among unvaccinated persons (*I*_*uv*_ [d^-1^]) was considered an experimental condition value in this study. Suppose *P*_*uv_ut*_ and *P*_*v_ut*_ are the proportions of asymptomatic infectors among unvaccinated and vaccinated spectators without any testing before events, respectively, and that they can be calculated on the basis of *I*_*uv*_ as in eqs. 1–4.

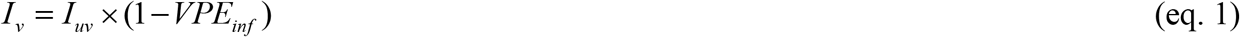

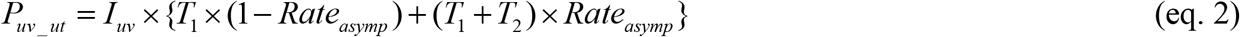

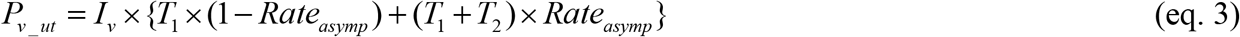

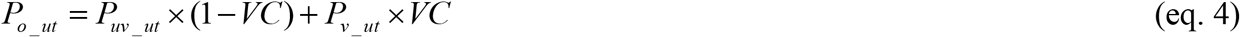

where *I*_*v*_ is the daily incidence of new infections among vaccinated persons [d^-1^], *VPE*_*inf*_ is the VPE against the risk of infection, *T*_*1*_ is the number of days between infectivity onset and symptom onset [d], *T*_*2*_ is the number of days between symptom onset and loss of infectivity [d], *Rate*_*asymp*_ is the ratio of the number of asymptomatic infected individuals to the number of all infected individuals, *P*_*o_ut*_ is the proportion of asymptomatic infectors among all spectators without any testing before events, and *VC* is the vaccination coverage.

We set the number of days between exposure and infectivity onsets (*T*_*0*_), *T*_*1*_, and *T*_*2*_ to 3, 2, and 7 days, respectively (He et al., 2020b). The total number of days of infectivity (*T*_*1*_ + *T*_*2*_) was the same (9 days), regardless of whether the individual was asymptomatic or symptomatic. *Rate*_*asymp*_ was set as an arithmetic mean of 0.460 and standard deviation of 0.141(He et al., 2020a) and followed a normal distribution left- and right-censored with 0 and 1, respectively.

The proportion of asymptomatic infectors among unvaccinated spectators who tested negative with polymerase chain reaction (PCR) testing 3 days prior to the event (*P*_*uv_pcr*_) was calculated as follows: First, the proportion of asymptomatic infectors or non-infectors among unvaccinated persons who had negative PCR results 3 days prior to the event was estimated as in eqs. 5–7. The subscript *i* indicates the number of days between exposure and the PCR test.

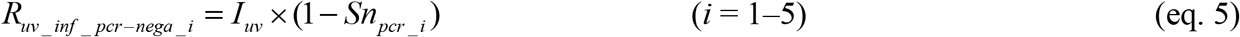

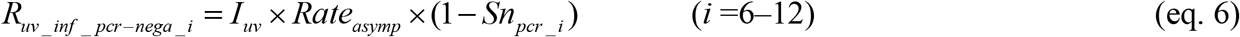

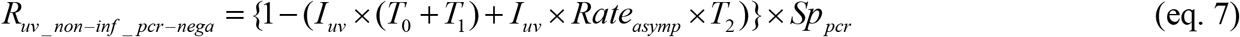

where *R*_*uv_inf_pcr-nega_i*_ is the proportion of asymptomatic infectors_*i*_ among unvaccinated persons who had negative PCR results 3 days prior to the event, *R*_*uv_non-inf_pcr-nega*_ is the proportion of non-infectors among unvaccinated persons who had negative PCR results 3 days prior to the event, and *Sn*_*pcr*_ and *Sp*_*pcr*_ are the sensitivity and specificity of the PCR test, respectively. The *Sn*_*pcr_i*_ was as follows (Kucirka et al., 2020): *i* = 1–3, 1-*Sp*_*pcr*_; *i* = 4, 0.33; *i* = 5, 0.62; *i* = 6–12: 0.8. The *Sp*_*pcr*_ value was 0.999.

The proportion of asymptomatic infectors among unvaccinated spectators with negative PCR results was then calculated as shown in eqs. 8–11.

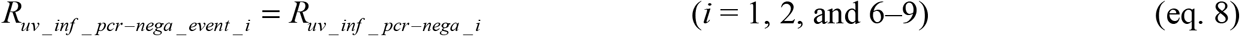

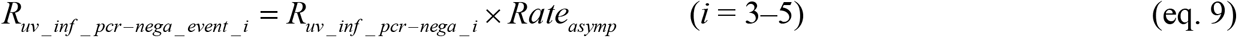

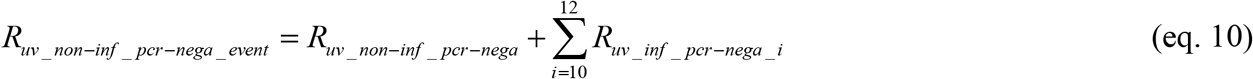

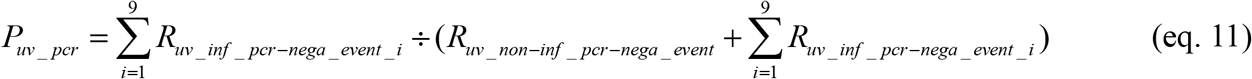

where *R*_*uv_inf_pcr_event_i*_ is the proportion of asymptomatic infectors_*i*_ at the time of the event among unvaccinated persons who had negative PCR test results, and *R*_*uv_non-inf_pcr_event*_ is the proportion of non-infectors at the time of event among unvaccinated persons who had negative PCR test results.

Similarly, the proportion of asymptomatic infectors among unvaccinated spectators who tested negative on the qualitative antigen test on the day of the event (*P*_*uv_ant*_) was calculated using eqs. 12–15.

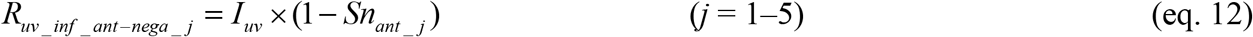

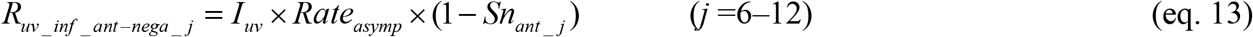

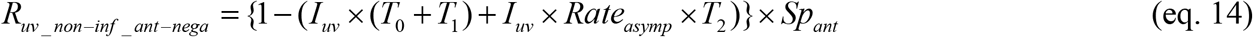

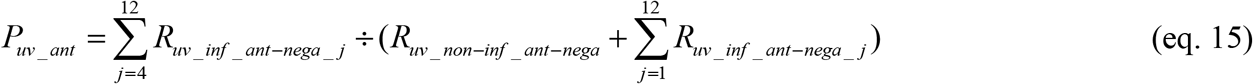

where *R*_*uv_inf_ant-nega_j*_ is the proportion of unvaccinated, asymptomatic infectors_*j*_ who had negative qualitative antigen test results on the day of the event, *R*_*uv_non-inf_ant-nega*_ is the proportion of unvaccinated, non-infectors who had negative qualitative antigen test results on the day of the event, *Sn*_*ant*_ and *Sp*_*ant*_ are the sensitivity and specificity of the qualitative antigen test, respectively, and *j* is the number of days between exposure and the qualitative antigen test.

The *Sn*_*ant_j*_ and *Sp*_*ant*_ were set as in eqs. 16-18 (Prince-Guerra et al., 2021).

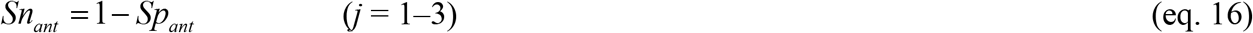

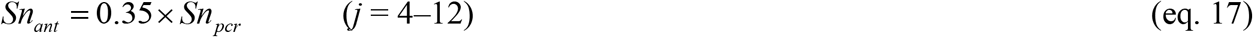

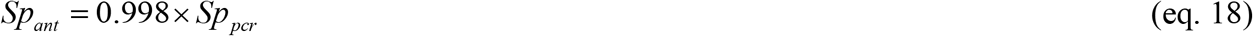

The proportion of vaccinated, asymptomatic infectors who tested negative with qualitative antigen testing on the day of the event (*P*_*v_ant*_) was calculated by replacing *I*_*uv*_ with *I*_*v*_ in the above equations.

In this study, *I*_*uv*_ was set to 10^−4^. The primary vaccines in Japan are the BNT162b2 vaccine (Pfizer-BioNTech) and mRNA-1273 (Moderna). An epidemiological study of COVID-19 cases between vaccinated and unvaccinated persons in Qatar showed that VPE against infection after the second dose of the BNT162b2 vaccine under the dominance of beta or delta variants was 70–78% within 3 months, 52% at 4 months, and 17-23% at 5-6 months, while VPE against severe, critical, or fatal cases of COVID-19 was reported to be maintained at ≥ 84% between 1 and 6 months (Chemaitelly et al., 2021). In addition, during the period when B.1.1.7 (alpha) or delta variants were predominant, VPE against infection with the mRNA-1273 vaccine was slightly higher than the BNT162b2 vaccine, while VPE against serious illness was similar between the two vaccines (Puranik et al., 2021). For the omicron variant, the VPE against symptomatic infections of the mRNA-1273 or BNT162b2 vaccines has ranged between 50–60% at 2–9 weeks after the second vaccination, 20–30% at 10–19 weeks, and 60–80% at 1–9 weeks after the third vaccination (UK Health Security Agency, 2021). The report also found that vaccination was more effective in preventing hospitalization. Therefore, in this study, the *VPE*_*inf*_ was set at 50% (corresponding to 4 months for beta or delta variants or 1–2 months for the omicron variant after the second dose of the vaccine), and the VPE against serious illness (*VPE*_*serious*_) was set at 80%. However, in Experiment B (see Section 2.2.), which aimed to evaluate the risk of infection and serious illness due to variations in *VPE*_*inf*_, we also analyzed the results under the condition that *VPE*_*inf*_ was 20% (corresponding to 5–6 months for beta or delta variants or 3–4 months for omicron variant after the second dose) or 80% (corresponding to 1–3 months for beta or delta variants after the second dose).

Second, the risk of infection to the spectators due to participation in mass gathering events was calculated as in eqs. 19–21, considering the *VPE*_*inf*_.

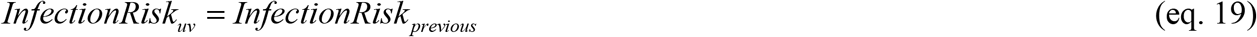

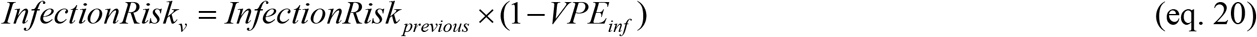

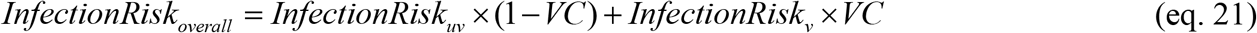

*InfectionRisk*_*uv*_, *InfectionRisk*_*v*_, and *InfectionRisk*_*overall*_ are risks of infection among unvaccinated, vaccinated, and all spectators due to participation in the event, respectively. *InfectionRisk*_*previous*_ is the risk of infection among spectators due to participation in the event in the dose-response model in previous studies (Murakami et al., 2021a; Yasutaka et al., 2021).

Third, in this study, in addition to the risk of infection, the risk of serious illness (i.e., cases of death or intensive care unit or ventilator treatment) was assessed as in eqs. 22–24.

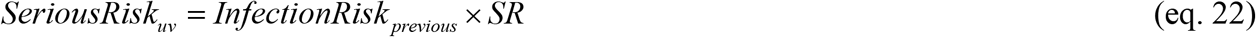

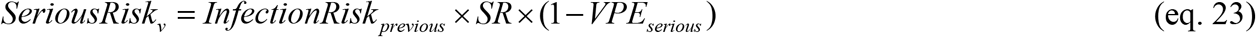

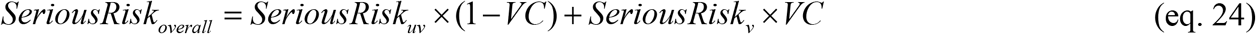

*SeriousRisk*_*uv*_, *SeriousRisk*_*v*_, and *SeriousRisk*_*overall*_ are risks of serious illness among unvaccinated, vaccinated, and overall spectators due to participation in the event, respectively. SR is the ratio of serious illness cases to the total number of infections (0.02) (Ministry of Health Labour and Welfare, 2021).

Fourth, regarding the effectiveness of unwoven masks, previous studies (Murakami et al., 2021a; Yasutaka et al., 2021) assumed that they could eliminate large particles of 95% (> 10 μm) but not small particles (< 10 μm). However, based on recent scientific findings, we assumed that small particles of 70% could be removed in this study (Ueki et al., 2020).

Fifth, to account for changes in infection risk due to the emergence of mutant variants, a sensitivity analysis was also performed under conditions where arithmetic means and standard deviations of the virus concentrations in saliva were increased by a factor of 10, 100, or 1000.

### 2.2. Experimental conditions

In this study, we analyzed the following three experiments A-C.

Experiment A was designed to compare the effectiveness of vaccinations, testing, and measures at stadiums in reducing the risk of infection and serious illness among spectators. Spectators were divided into the following 10 conditions: (1) unvaccinated, no testing, no measures; (2) unvaccinated, PCR test 3 days prior to the event, no measures; (3) unvaccinated, qualitative antigen test on the day of the event, no measures; (4) vaccinated, no testing, no measures; (5) vaccinated, qualitative antigen test on day of the event, no measures; (6) unvaccinated, no testing, with measures; (7) unvaccinated, PCR test 3 days prior to the event, with measures; (8) unvaccinated, qualitative antigen test on the day of the event, with measures; (9) vaccinated, no testing, with measures; (10) vaccinated, qualitative antigen test on the day of the event, with measures.

Experiment B was conducted to determine how differences in *VPE*_*inf*_ led to differences in the risk of infection and serious illness under different *VCs*. The risk of infection and serious illness was estimated under the following conditions: *VPE*_*inf*_ was set at 20%, 50%, and 80%; *VC* was set at 0%, 10%, 20%, 30%, 40%, 50%, 60%, 70%, 80%, 90%, and 100%. We considered the condition that spectators were not tested and measures were taken. The conditions of *VC* of 0% or 100% and *VPE*_*inf*_ of 50% were the same as those in Experiment A. The VC in Japan was 78% as of December 11, 2021 (Our World in Data, 2021).

Experiment C was conducted to compare the differences in the number of newly infected individuals and those with serious illness with and without zoning. With zoning, both VT seats and regular seats had 20,000 spectators, and the number of spectators without zoning was 40,000. All spectators in VT seats were assumed to be vaccinated without testing. An on-site investigation of professional sports in Japan reported that the majority of people in VT seats (98.2-99.0% in two baseball events) had proof of vaccination (Nippon Professional Baseball, 2021). Regular seats were used for spectators without confirmation by the event organizer regarding vaccination status or proof of negative test results, and some of these spectators may have included vaccinated persons.

The conditions without zoning were *VC* of 0%, 10%, 20%, 30%, 40%, 50%, 60%, 70%, 80%, 90%, and 100% (same as Experiment B). The zoning conditions were *VC* of 50% (100% for VT seats and 0% for regular seats), 60% (100% for VT seats and 20% for regular seats), 70% (100% for VT seats and 40% for regular seats), 80% (100% for VT seats and 60% for regular seats), and 90% (100% for VT seats and 80% for regular seats).

Furthermore, in a sensitivity analysis to account for differences in seat ratios, the zoning conditions in which the number of spectators for VT package seats and regular seats were changed as follows: VT package seats of 4,000 + regular seats of 36,000 (*VC* of 10%), 8,000 + 32,000 (20%), 12,000 + 28.000 (30%), 16,000 + 24,000 (40%), 24,000 + 16,000 (60%), 28,000 + 12,000 (70%), 32,000 + 8,000 (80%), and 36,000 + 4,000 (90 %). Here, the *VC* for regular seats was set to 0%.

The number of newly infected individuals or those with serious illness with zoning in place was considered as the sum of those in the simulations, when the environmental exposure models were run separately for VT seats only and regular seats only.

We ran 10,000 Monte Carlo simulations for each scenario.

### 2.3. Uncertainties and interpretive notes

There were some uncertainties and interpretive notes in this study. Some have been described in detail in previous reports (Murakami et al., 2021a; Yasutaka et al., 2021). First, the parameters were set based on the knowledge currently available, and the validity of the model results was verified based on case studies of Japanese professional baseball and football events in the fiscal year 2020 (Yasutaka et al., 2021). However, some factors may not be accounted for, such as differences in the loudness of voice and the inactivation of the virus on finger and hair surfaces. Second, we assumed viral emissions and exposures from talking, coughing, and sneezing in the concourse, restrooms, concession areas, and stands, but we did not cover infection risk assessment outside of the scenarios we set up, such as group conversations associated with eating and drinking in the concourse. Similarly, the risk of infection associated with the movement of people, eating, and drinking outside the stadium was not assessed.

The VPE used in this study was based on data from individuals vaccinated with the BNT162b2 vaccines in Qatar during periods when beta and delta variants were prevalent (Chemaitelly et al., 2021). Because this previous study was not conducted in a randomized control trial design, infection control behaviors might differ between vaccinated and unvaccinated individuals, which could underestimate VPE and overestimate the risk of infection and serious illness.

SR was set at 0.02 based on values for Japan as a whole, but it is known that SR is higher among the elderly (Ministry of Health Labour and Welfare, 2021). If spectators attending mass gathering events have a higher proportion of young people than in Japan as a whole, the risk of serious illness may be overestimated.

Furthermore, there are uncertainties in this study that have not previously been reported in the estimation of the number of newly infected individuals and those with serious illness when VT seats and regular seats are zoned. In this study, environmental exposure model simulations were conducted separately for VT seats and regular seats, but in reality, VT seats and regular seats are not in separate stadiums, but in one stadium. When environmental exposure model simulations were performed separately, the virus concentrations in air differed from what would be expected if a single stadium was considered. This caused the differences in the number of newly infected individuals or those with serious illness among the five types of spectators, particularly in the fifth category (i.e., “others”). However, in Experiment C, the number of newly infected individuals or those with serious illness in the fifth category of spectators (i.e., “others”) was less than 0.1% the total number of newly infected individuals or those with serious illness. Therefore, the difference found in this analysis was negligible and was not expected to affect the results. In addition, it was assumed that there was no difference in spectator behavior (e.g., mask wearing and cheering) between VT package seats and regular seats. This assumption is valid as on-site surveys conducted from October to November 2021 revealed that there was no difference in spectator behavior between users and non-users of the VT package at mass gathering events, including professional football games (Subcommittee on Novel Coronavirus Disease Control, 2021).

## 3. Results

### 3.1. Risk of infection and serious illness among spectators with and without vaccination, testing, and measures in place (Experiment A)

Without measures in place at stadiums, the average risk of infection for unvaccinated, non-tested spectators was 5.2×10^−4^, whereas those with PCR tests 3 days prior and qualitative antigen tests on day of the event had an average risk of infection of 3.2×10^−4^ and 4.0×10^−4^, respectively, showing a 37% and 24% reduction in the risk of infection (Figure 1a). Similarly, with measures in place, the risk reduction effects of the PCR and qualitative antigen tests were 39% and 24%, respectively. For vaccinated spectators, the risk reduction effect of the qualitative antigen test was 24% in the absence of measures and 25% in the presence of measures.

**Figure 1.**
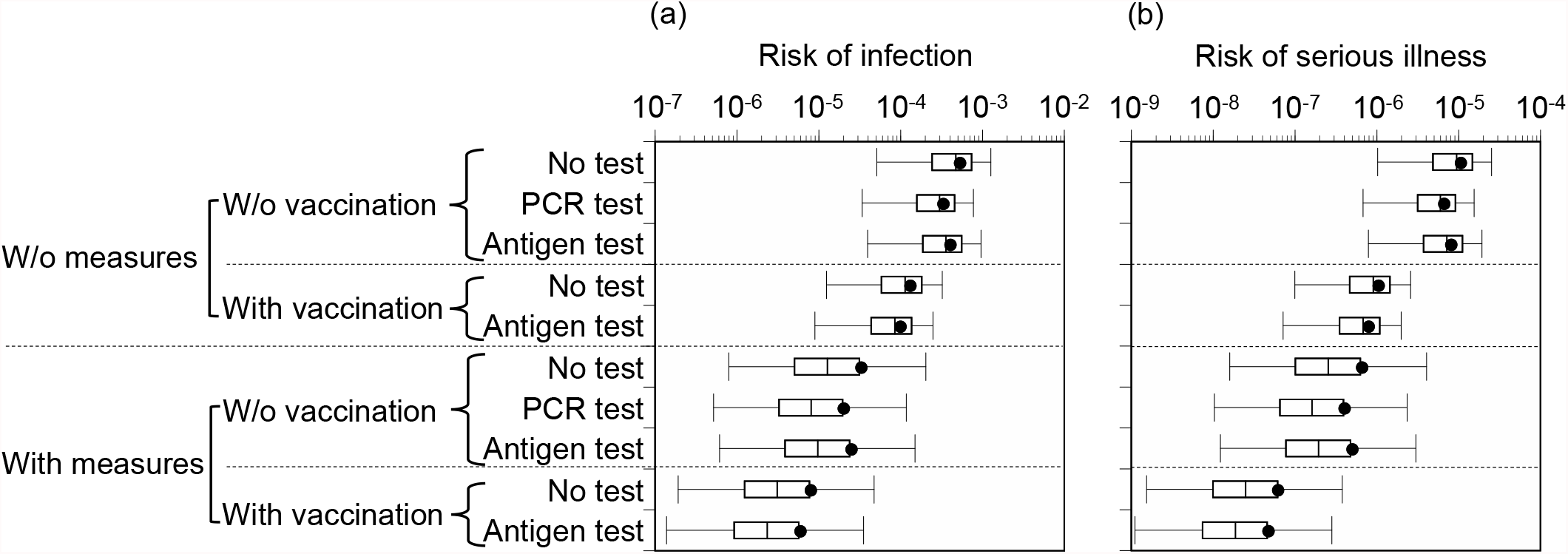
Comparison of risk between with and without vaccination, testing, and measures. (a) Risk of infection, (b) risk of serious illness. Box-and-whisker plots represent 2.5, 25, 50, 75, and 97.5 percentiles. Closed circles represent average values (arithmetic mean) of simulations. PCR test: polymerase chain reaction test 3 days prior to the event. Antigen test: qualitative antigen test on day of the event.

Under the condition that *VPE*_*inf*_ was 50%, the risk of infection among vaccinated spectators without testing and measures in place was 1.3×10^−4^, indicating that vaccinations had a 75% risk reduction effect. Similarly, the risk reduction due to vaccination was 76 % under the condition of no testing and with measures. The risk of serious illness in unvaccinated spectators without testing and measures in place was 1.0×10^−5^, while that in vaccinated spectators was 1.0×10^−6^, which represented a 90% reduction in the risk of serious illness (Figure 1b). Similarly, there was a 90% reduction in the risk of serious illness under the same conditions with measures in place. Importantly, under the condition where measures were in place, the risk of infection and serious illness was lower among non-tested, vaccinated spectators than that in unvaccinated spectators with PCR and qualitative antigen tests. The ratios of vaccinated spectators without testing to unvaccinated spectators with testing were 0.32–0.40 for risk of infection and 0.13–0.16 for risk of serious illness, respectively.

The risk of infection and serious illness among vaccinated spectators without any testing and measures in place was 1.3×10^−4^ and 1.0×10^−6^, respectively, whereas it was 3.2×10^−5^ and 6.4×10^−7^, respectively, in unvaccinated spectators without testing and with measures in place. The risk of infection and serious illness among vaccinated spectators without measures was 4.0 and 1.6 times higher than that among unvaccinated spectators with measures, respectively. Similarly, the risks of both infection and serious illness were 16 times higher among vaccinated spectators without any testing and measures in place compared with vaccinated spectators without testing but with measures in place.

Even under the assumption of mutant variants existing (10–1000 times of virus concentrations), the sensitivity analysis confirmed that the risk of infection and serious illness was lower in non-tested vaccinated spectators than in tested unvaccinated spectators and that the risk of infection was lower among unvaccinated spectators with measures than among vaccinated spectators without measures, irrespective of the virus concentration conditions (Figure S1).

### 3.2. Effect of VPE and VC on risk of infection (Experiment B)

We quantitatively analyzed the risk of infection and serious illness when *VC* ranged from 0% to 100% and *VPE*_*inf*_ was 20%, 50%, and 80% (Figure 2a, b). When *VC* increased from 0% to 80%, the risk of infection decreased to 0.12 times with *VPE*_*inf*_ of 80%; however, it decreased to 0.35 times with *VPE*_*inf*_ of 50% and 0.70 times with *VPE*_*inf*_ of 20%. The risk of serious illness was 0.12, 0.21, and 0.30 times, respectively. The risk of infection with *VPE*_*inf*_ of 20% and *VC* of 80% were similar to those with *VPE*_*inf*_ of 80% and *VC* of 20%. Sensitivity analysis with different virus concentrations also showed similar results (Figure S2).

**Figure 2.**
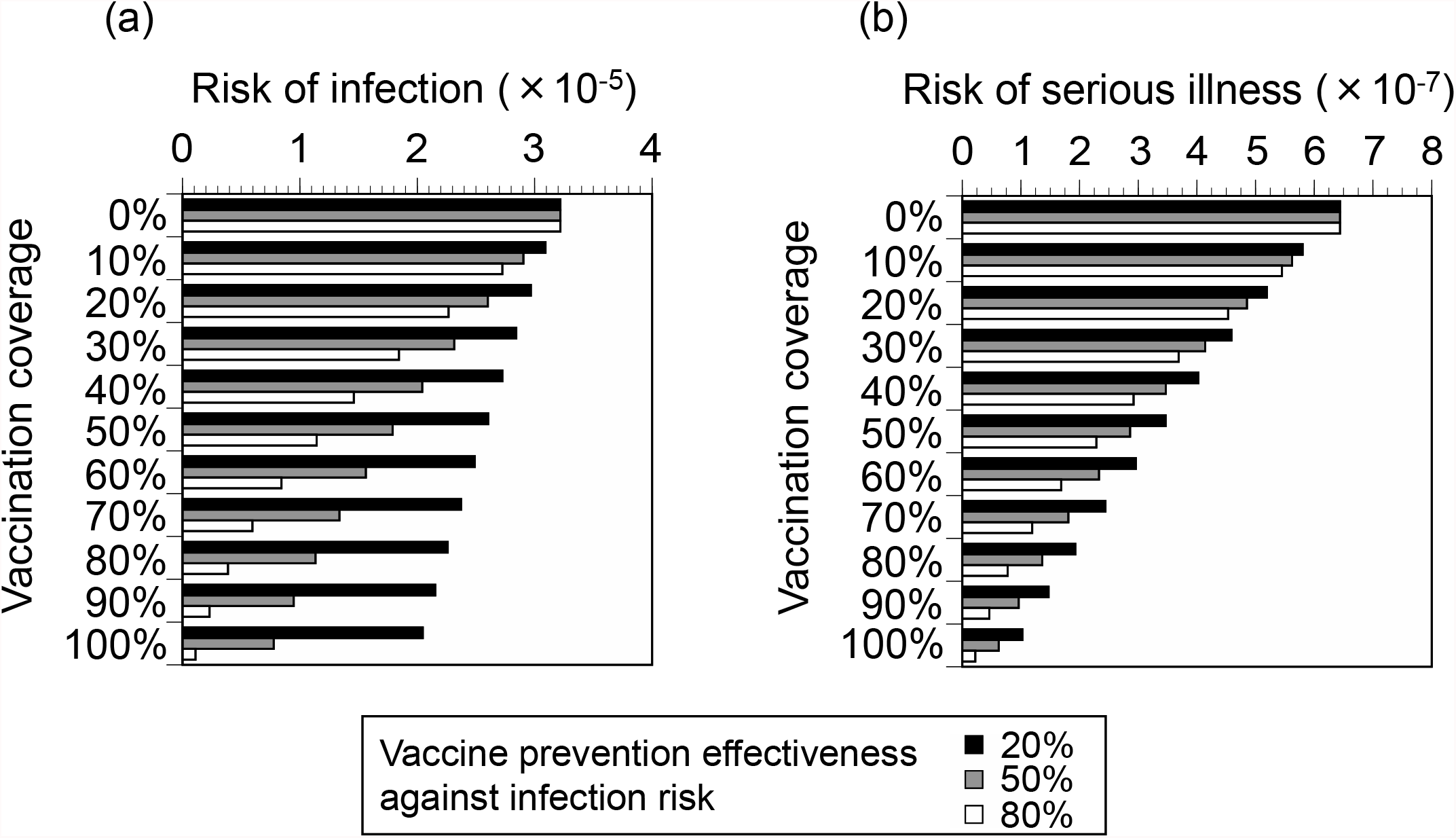
Differences in risk due to variations in vaccination coverages and vaccine prevention effectiveness against infection. (a) Risk of infection, (b) risk of serious illness.

### 3.3. Risk of infection risk with and without seat zoning (Experiment C)

Figures 3a shows the number of newly infected individuals with and without VT and regular seat zoning. When the overall *VC* was 50%, the total number of newly infected individuals was 0.72 in the absence of zoning and 0.76 in the presence of zoning; the total number of newly infected individuals without zoning was slightly lower than that with zoning. Of these, the number of newly infected individuals among vaccinated and unvaccinated spectators without zoning was 0.24 and 0.48, respectively, while those with zoning were 0.14 and 0.62, respectively. The risk of infection among vaccinated spectators was lower in the presence of zoning, whereas it was lower in the absence of zoning among unvaccinated spectators. The same was true for the number of infected individuals with serious illness (Figure 3b).

**Figure 3.**
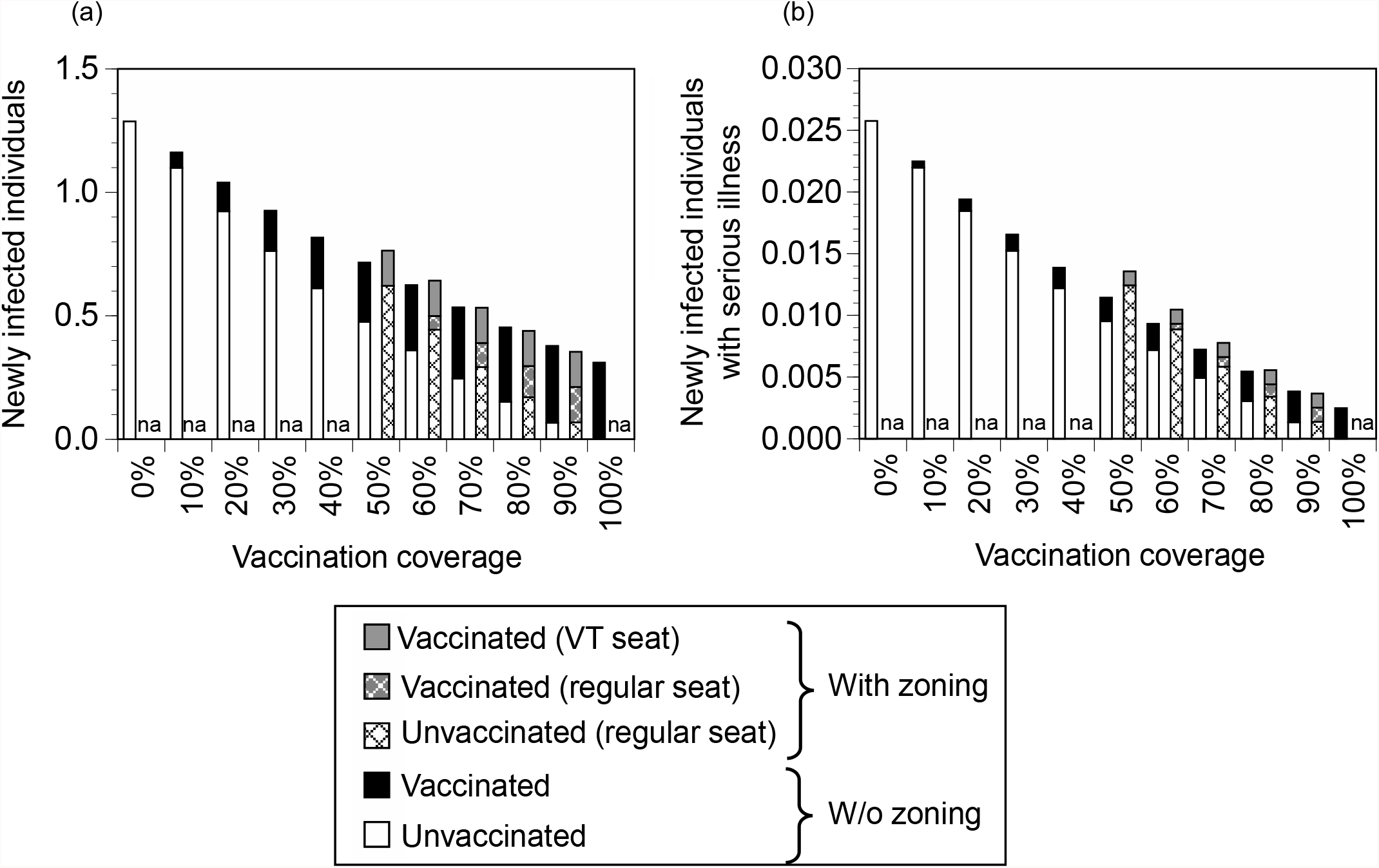
Differences in risk due to seat zoning. (a) The number of newly infected individuals, (b) the number of newly infected individuals with serious illness. VT seats: seats for users of vaccine-testing packages.

However, when the overall *VC* was 80%, the differences in the number of newly infected individuals among overall spectators, vaccinated spectators, and unvaccinated spectators in the presence or absence of zoning were within a factor of only 1.03, 1.12, and 1.13, respectively. Similarly, in the number of newly infected individuals with serious illness, the differences between with and without zoning were negligibly small when the overall *VC* exceeded 80%.

Sensitivity analysis with different virus concentrations (Figure S3) and different ratios of spectators between VT package and regular seats (Figure S4) confirmed that the total number of newly infected individuals or those with serious illness was lower in the absence of zoning than in the presence of zoning with an overall *VC* of 50% and that the difference in the number of newly infected individuals or those with serious illnesses between those with and without zoning was negligibly small when the overall *VC* exceeded 80%.

## 4. Discussion

In this study, we quantitatively assessed the risk of infection and serious illness by using an environmental exposure model to discuss the effects of introducing VT packages to mass gathering events.

First, we evaluated the effects of vaccination, testing, and implementation of other measures at stadiums, such as wearing masks and ventilation, in reducing the risk of infection and serious illness among spectators. Testing unvaccinated spectators produced an approximately 20–40% reduction in the risk of infection. The effects of vaccinations, assuming *a VPE*_*inf*_ of 50%, was a 75% reduction in the risk of infection. Furthermore, the reduction in the risk of serious illness was even greater with vaccination than with testing of unvaccinated spectators. Importantly, the risk of infection among vaccinated spectators in the absence of measures was 4.0 and 16 times higher than that among unvaccinated spectators with measures and vaccinated spectators with measures in place. VPE against infection with the beta or delta variants was approximately 70–80% within 3 months after the second dose of the BNT162b2 vaccine, 50% after 4 months, and 20% after 5–6 months (Chemaitelly et al., 2021). In fact, even though the United Kingdom had more than 50% of its population vaccinated with two doses of the vaccine at the UEFA Euro 2020 Final on July 11, 2021 (Our World in Data, 2021), the lack of adequate measures, such as mask wearing, resulted in more than 3,000 newly infected individuals after the emergence of delta variant. The omicron variant further reduces the VPE against symptomatic infection by 50–60% at 2–9 weeks and 20–30% at 10–19 weeks after the second dose of the vaccine (UK Health Security Agency, 2021). Since the VPE against infection decreases, it is important for vaccinated spectators and organizers to take measures, such as mask wearing and proper ventilation, to reduce the risk of infection at mass gathering events, even if VC is sufficiently high. While it has been estimated that wearing a mask is particularly effective in reducing the risk of infection (Yasutaka et al., 2021), the proportion of mask wearing during the game at mass gathering events varies from country to country. In Japan, the proportion of mask-wearing football spectators was ≥ 93% (Subcommittee on Novel Coronavirus Disease Control, 2021), whereas in the United Kingdom, it was recently reported at < 30%, despite the requirement to wear masks (The United Kingdom Government, 2021b).

Next, we evaluated the effects of VC and VPE on the risk of infection. Increasing *VC* from 0% to 80% greatly reduced to 0.12 times of the risk of infection with a *VPE*_*inf*_ of 80%, but only reduced it by 0.70 times with a *VPE*_*inf*_ of 20%. Furthermore, the risk of infection with a *VC* of 80% and *VPE*_*inf*_ of 20% were similar to those with a *VC* of 20% and *VPE*_*inf*_ of 80%. Although the increase in VC in Japan was one of the highest in the world, it still took five months to improve from a two-dose VC of 20% (as of July 11, 2021) to 78% (December 11, 2021) (Our World in Data, 2021). Considering the changes in VPE, if it takes more than 5 months to increase the national VC from 20% to 80%, it is unlikely to be effective in reducing the risk of infection among spectators at mass gathering events. It remains to be seen to what extent and how long VPE against risk of infection will be maintained after 3 doses of vaccine in the face of emerging mutant variants such as omicron. It may be unproven that simply boosting the current vaccine will be enough to maintain a high level of VPE against infection for the entire population for a long time. Selective vaccination strategies for people whose antibody levels are likely to decline and the development of vaccines that maintain long-term VPE against infection are expected.

In addition, the role of zoning of VT and regular seats was evaluated for the number of newly infected individuals and those with serious illness among spectators. When the overall *VC* was 50%, the number of newly infected individuals without zoning was lower in overall spectators and unvaccinated spectators than in those with zoning. This is because a reduction in the risk of infection occurs when vaccinated and unvaccinated spectators are seated next to each other. However, with a *VC* of 80% or more, there was a negligible difference in the number of newly infected individuals or those with serious infections with or without zoning. These results indicated that there may be little need to implement seat zoning in terms of infection risk reduction. It would be worthwhile to examine the benefits and disadvantages of zoning mass gathering events from a practical viewpoint (e.g., ease of selling and arranging seats) and in terms of the demands of spectators (e.g., requests for VT seats from vaccinated spectators), rather than the risk of infection.

## 5. Conclusions

In this study, we discussed the effects of introducing the VT package by quantitatively assessing the risk of infection and serious illness among spectators at mass gathering events using an environmental exposure model. The main findings are as follows.

- If VPE against infection was 50% (corresponding to 4 months for beta or delta variants or 1–2 months for the omicron variant after the second dose of the vaccine), the risk of infection among vaccinated spectators without taking measures at stadiums, such as mask wearing, was higher than that among unvaccinated spectators who took measures. Even if vaccinated, thoroughly implemented individual measures at stadiums are important in reducing the risk of infection during mass gathering events.
- The risk of infection with *VC* of 80% and *VPE*_*inf*_ of 20% (corresponding to 5–6 months for beta or delta variants or 3–4 months for omicron variant after the second dose of the vaccine) was similar to that with a *VC* of 20% and *VPE*_*inf*_ of 80% (corresponding to 1–3 months for beta or delta variants after the second dose). Maintaining a high level of VPE against infection is important for reducing the risk of infection. Since it often takes more than 5 months to improve vaccine coverage, it is necessary to develop vaccines that maintain long-term VPE against infection, as well as vaccination strategies that selectively vaccinate individuals whose antibody levels are likely decline.
- There was a negligible difference in the risk of infection between zoning VT seats and regular seats with a *VC* of 80% or more. Even under conditions of lower *VC*, the risks of infection without zoning were lower in overall spectators and unvaccinated spectators. There may be little need to implement seat zoning for infection risk reduction.

## Data Availability

All data produced in the present work are contained in the manuscript.

## Authorship contribution statement

**Michio Murakami:** Conceptualization; Data curation; Methodology; Visualization; Project administration; Writing –original draft

**Tsukasa Fujita**: Formal analysis; Methodology; Writing –review & editing

**Yuichi Iwasaki**: Formal analysis; Methodology; Writing –review & editing

**Masaki Onishi**: Writing –review & editing

**Wataru Naito**: Writing –review & editing

**Seiya Imoto**: Project administration; Supervision; Writing –review & editing

**Tetsuo Yasutaka**: Conceptualization; Methodology; Project administration; Writing –review & editing

## Competing interests

### Support for the reported work

This research received no external financial or non-financial support.

### Relevant support outside this work

Y.I., M.O., W.N., and T.Y. report a relationship with Kao Corporation: funding grants.

M.O., W.N., and T.Y. report a relationship with Nippon Professional Baseball Organization, Yomiuri Giants, Tokyo Yakult Swallows, the Japan Professional Football League, and the Japan Professional Basketball League: funding grants.

M.O. reports a relationship with Kashima Antlers FC: funding grants.

### Intellectual property

There are no patents to disclose.

### Other activities

This study was conducted as part of a comprehensive research project, comprising members from two private companies, Kao Corporation and NVIDIA Corporation, Japan. No authors in this study belong to these companies. M.M., M.O., W.N, S.I., and T.Y. attended the new coronavirus countermeasures liaison council jointly established by the Nippon Professional Baseball Organization and Japan Professional Football League as experts without any rewards. M.O., W.N., and T.Y. are advisors to the Japan National Stadium and Japan Professional Football League. Other authors declare no competing interests. The findings and conclusions of this article are solely the responsibility of the authors and do not represent the official views of any institution.

## Acknowledgements

We would like to thank Editage (www.editage.com) for English language editing. No external financial support is used for this article.

## Appendix

**Figure S1.**
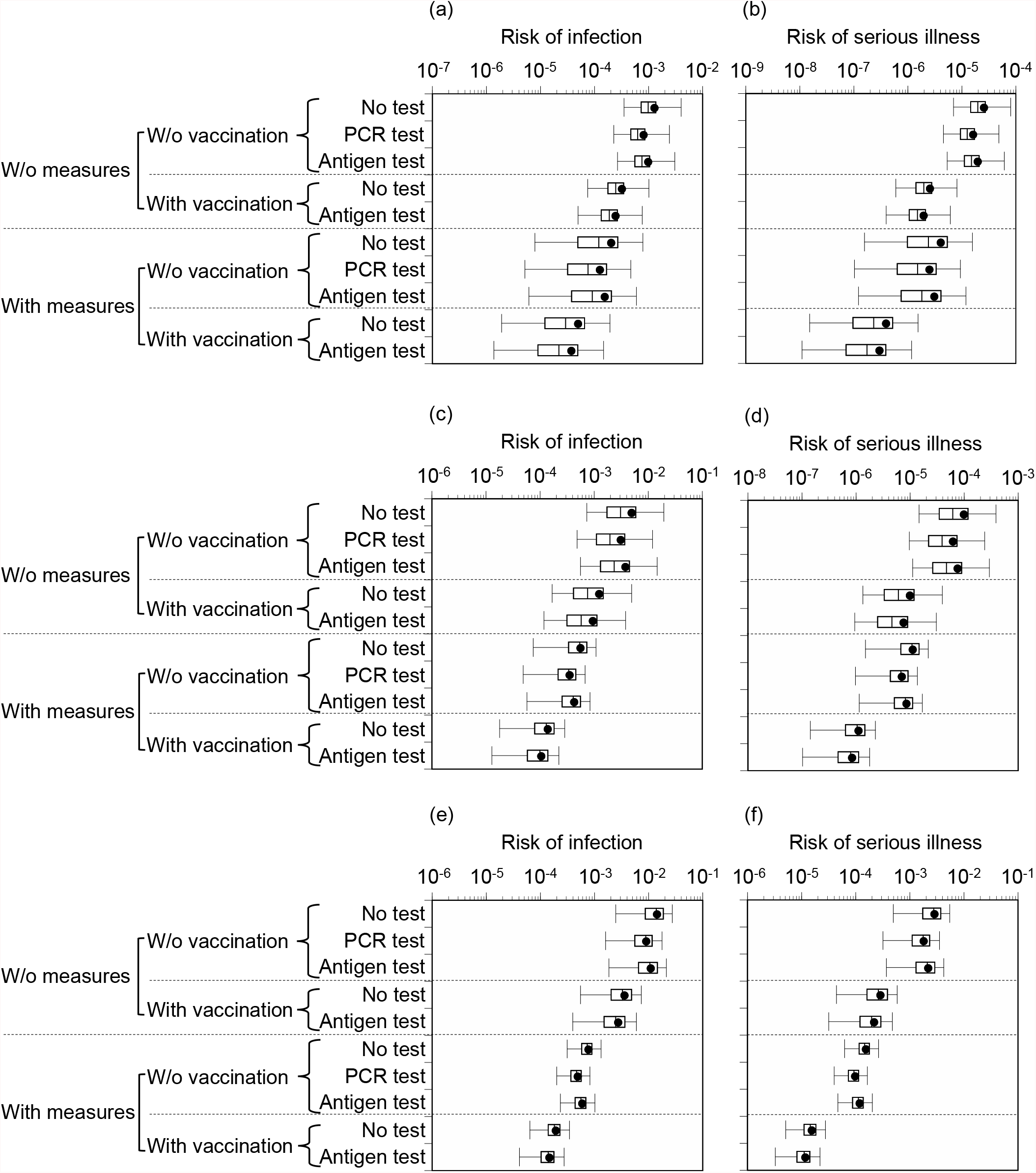
Comparison of risk between with and without vaccination, testing, and measures. (a) Risk of infection (virus concentration: ×10 times), (b) risk of serious illness (virus concentration: ×10 times), (c) risk of infection (virus concentration: ×100 times), (d) risk of serious illness (virus concentration: ×100 times), (e) risk of infection (virus concentration: ×1000 times), (f) risk of serious illness (virus concentration: ×1000 times). Box-and-whisker plots represent 2.5, 25, 50, 75, and 97.5 percentiles. Closed circles represent average values (arithmetic mean) of simulations. PCR test: polymerase chain reaction test 3 days prior to the event. Antigen test: qualitative antigen test on day of the event.

**Figure S2.**
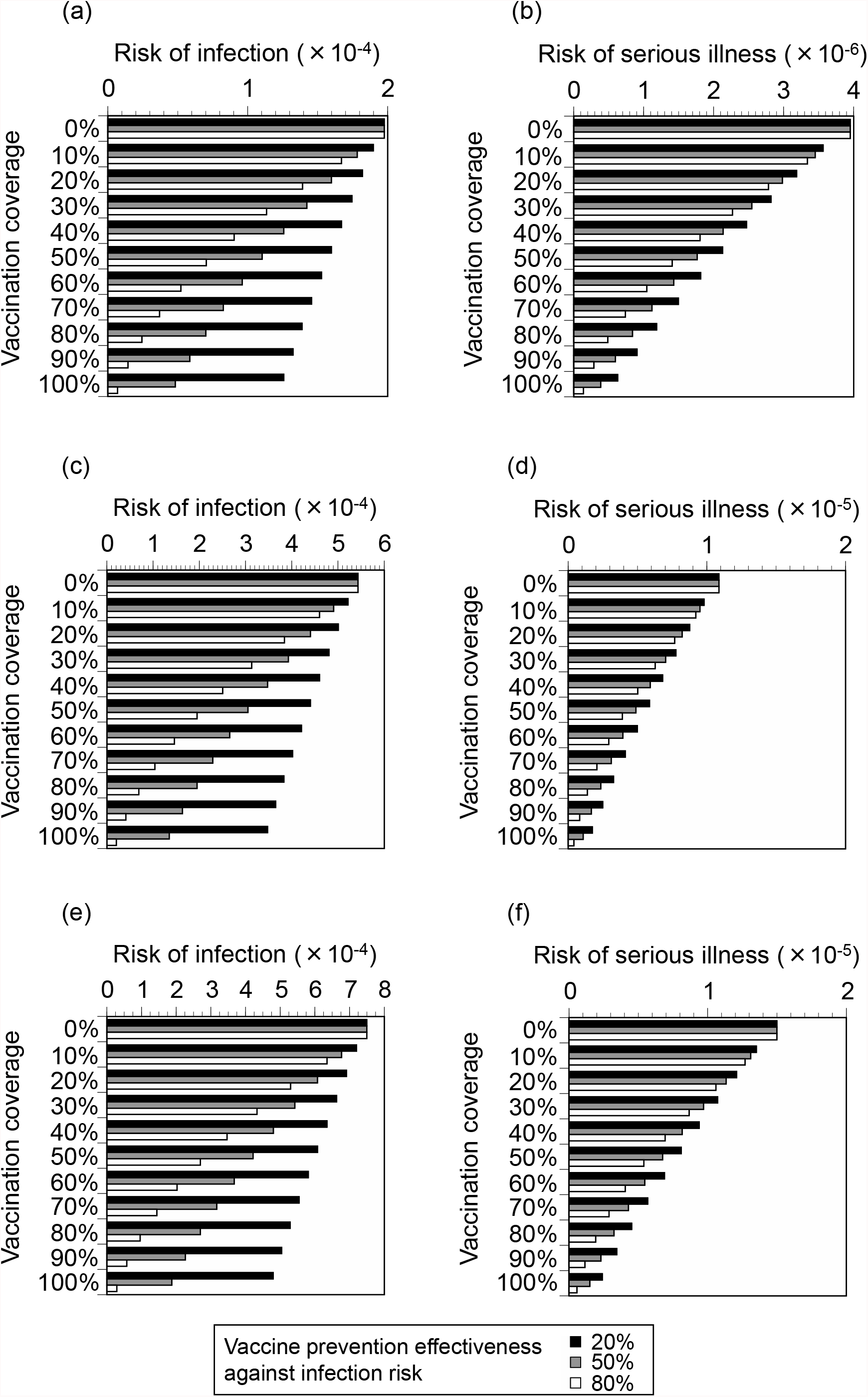
Differences in risk due to variations in vaccination coverages and vaccine prevention effectiveness against infection. (a) Risk of infection (virus concentration: ×10 times), (b) risk of serious illness (virus concentration: ×10 times), (c) risk of infection (virus concentration: ×100 times), (d) risk of serious illness (virus concentration: ×100 times), (e) risk of infection (virus concentration: ×1000 times), (f) risk of serious illness (virus concentration: ×1000 times).

**Figure S3.**
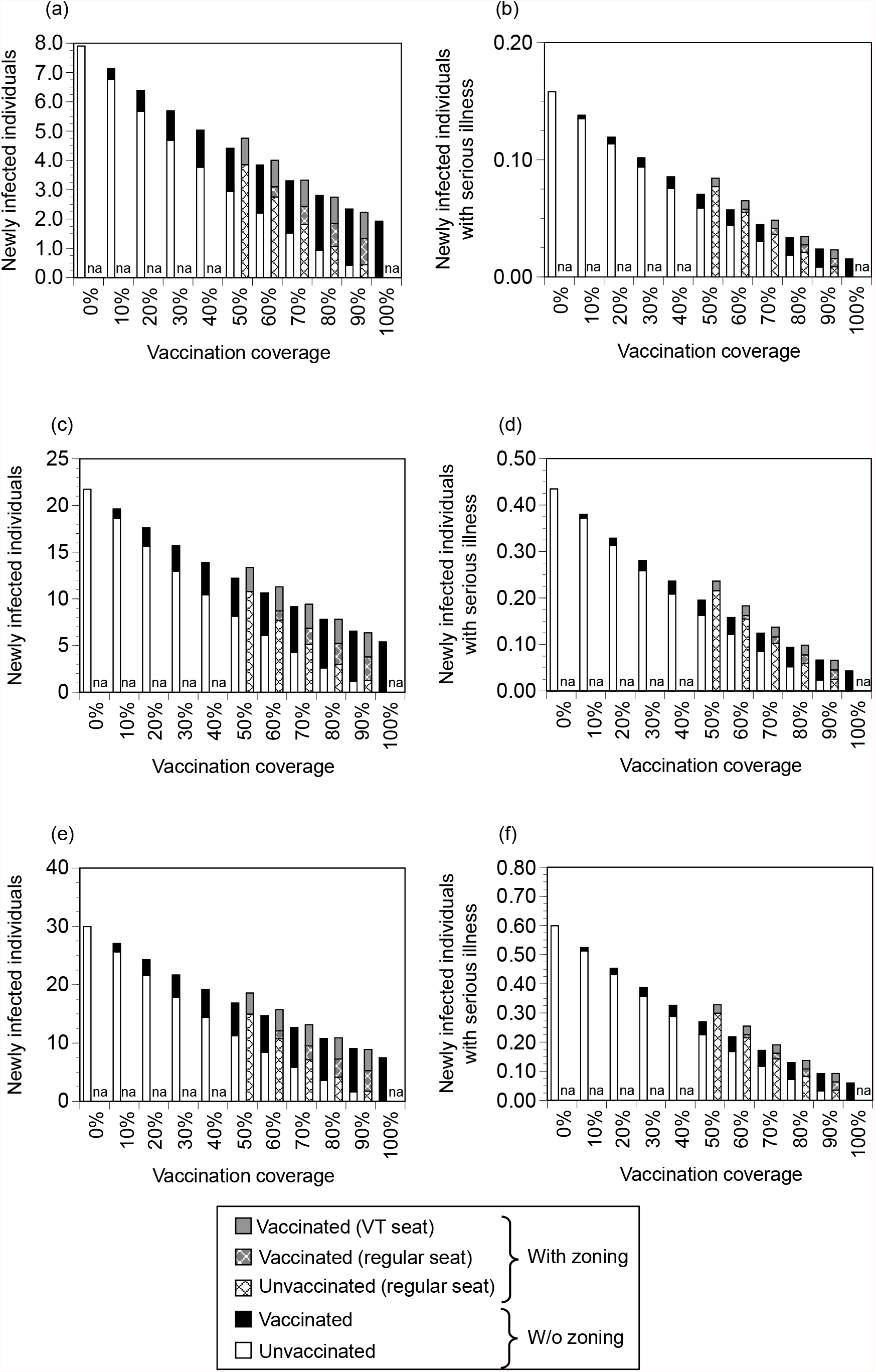
Differences in risk due to seat zoning. (a) The number of newly infected individuals (virus concentration: ×10 times), (b) the number of newly infected individuals with serious illness (virus concentration: ×10 times), (c) the number of newly infected individuals (virus concentration: ×100 times), (d) the number of newly infected individuals with serious illness (virus concentration: ×100 times), (e) the number of newly infected individuals (virus concentration: ×1000 times), (f) the number of newly infected individuals with serious illness (virus concentration: ×1000 times). VT seats: seats for users of vaccine-testing packages.

**Figure S4.**
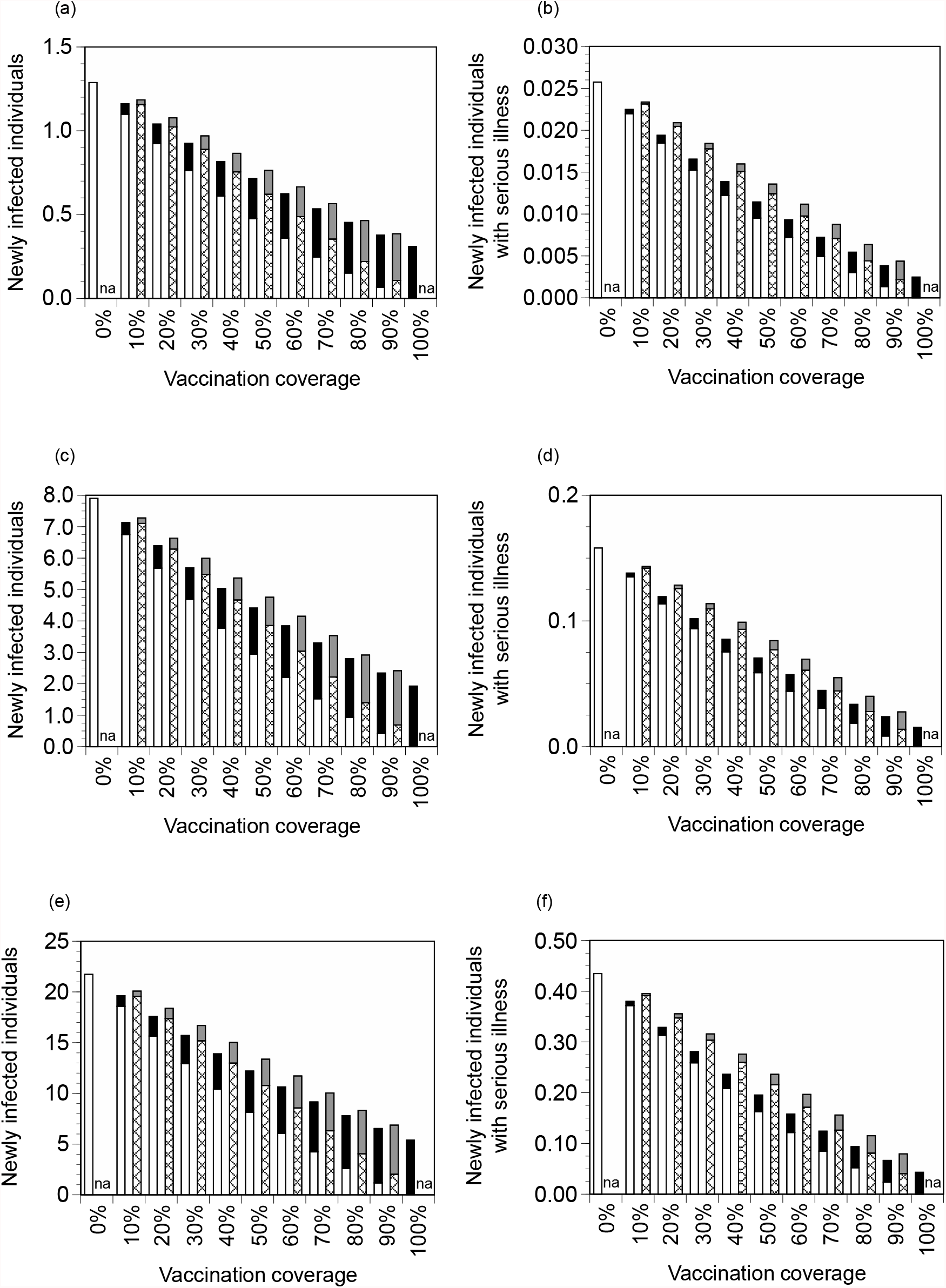
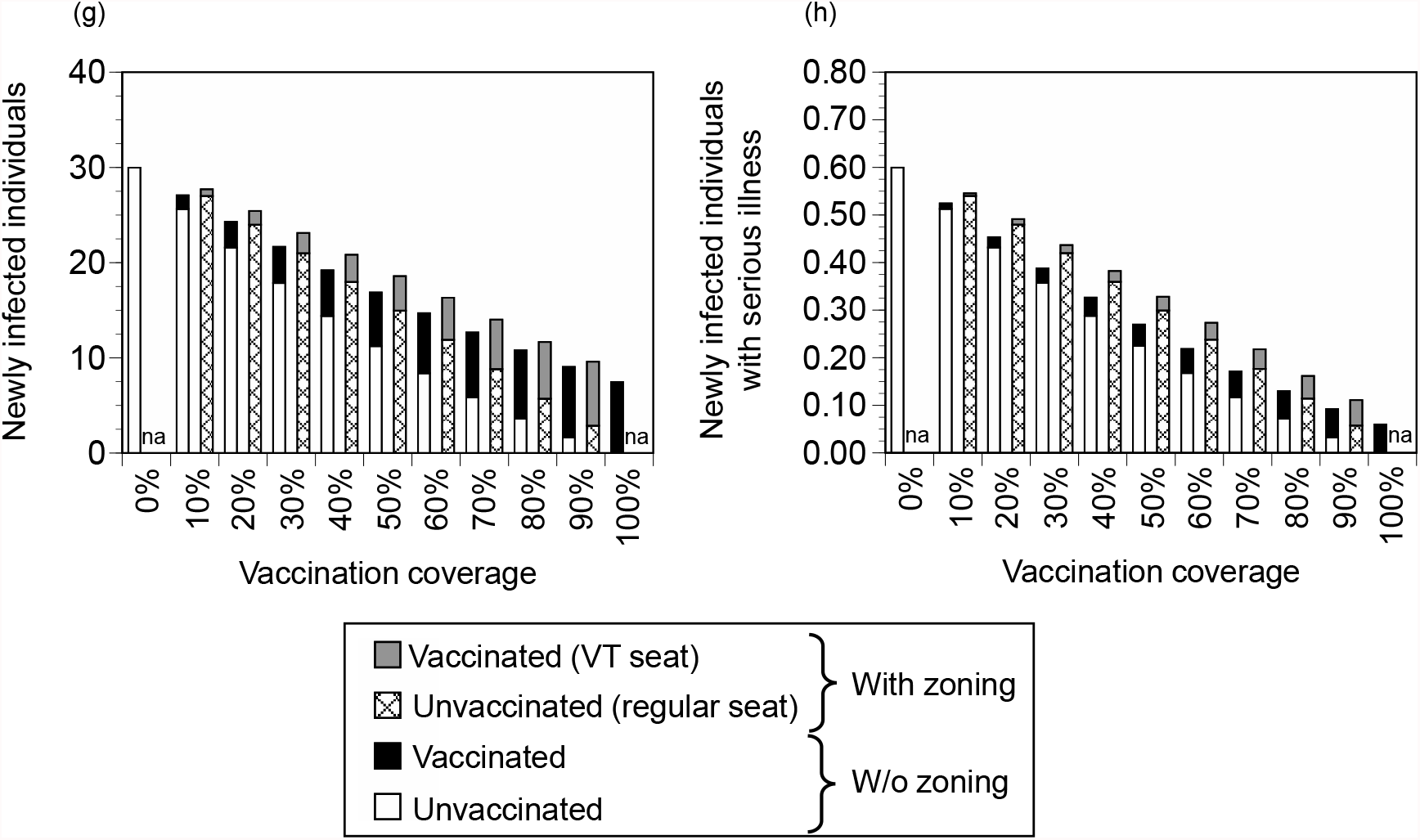
Differences in risk due to zoning when the numbers of spectators in VT seats and regular seats are varied. All spectators in VT seats are vaccinated; all spectators in regular seats are unvaccinated. Vaccination coverage is equal to the ratio of VT spectators to all spectators. (a) The number of newly infected individuals (virus concentration: ×1 times), (b) the number of newly infected individuals with serious illness (virus concentration: ×1 times), (c) the number of newly infected individuals (virus concentration: ×10 times), (d) the number of newly infected individuals with serious illness (virus concentration: ×10 times), (e) the number of newly infected individuals (virus concentration: ×100 times), (f) the number of newly infected individuals with serious illness (virus concentration: ×100 times). (g) the number of newly infected individuals (virus concentration: ×1000 times), (h) the number of newly infected individuals with serious illness (virus concentration: ×1000 times). VT seats: seats for users of vaccine-testing packages.

## References

Advisory Committee on the Basic Action Policy, 2022. The 20th meeting materials. https://www.cas.go.jp/jp/seisaku/ful/taisakusuisin/taisyo/dai20/gijishidai.pdf. (accessed 20 January, 2022). (in Japanese)

BBC, 2021. Covid: Germany puts major restrictions on unvaccinated. https://www.bbc.com/news/world-europe-59502180. (accessed 9 December 2021).

Cabinet Secretariat, 2021. Restoration of daily life to support the stability and security of citizen’s work and life. https://corona.go.jp/package/. (accessed 9 December, 2021). (in Japanese)

Chemaitelly, H., et al., 2021. Waning of BNT162b2 vaccine protection against SARS-CoV-2 infection in Qatar. N. Eng. J. Med. 385, e83.

Chiampas, G. T., Ibiebele, A. L., 2021. A sports practitioner’s perspective on the return to play during the early months of the COVID-19 pandemic: Lessons learned and next steps. Sports Med. 51, 89–96.

Desson, Z., et al., 2021. Finding the way forward: COVID-19 vaccination progress in Germany, Austria and Switzerland. Health Policy Technol., 100584.

He, W., et al., 2020a. Estimation of the basic reproduction number, average incubation time, asymptomatic infection rate, and case fatality rate for COVID-19: Meta-analysis and sensitivity analysis. J. Med. Virol. 92, 2543–2550.

He, X., et al., 2020b. Temporal dynamics in viral shedding and transmissibility of COVID-19. Nat. Med. 26, 672–675.

Kucirka, L. M., et al., 2020. Variation in false-negative rate of reverse transcriptase polymerase chain reaction–based SARS-CoV-2 tests by time since exposure. Ann. Intern. Med. 173, 262–267.

Levin, E. G., et al., 2021. Waning immune humoral response to BNT162b2 Covid-19 vaccine over 6 months. N. Eng. J. Med. 385, e84.

Ministry of Health Labour and Welfare, 2021. Current elleven knowledge about COVID-19 (December 2021). https://www.mhlw.go.jp/content/000788485.pdf. (accessed 9 December 2021). (in Japanese)

Murakami, M., et al., 2021a. COVID-19 risk assessment at the opening ceremony of the Tokyo 2020 Olympic Games. Microb. Risk Anal., 100162.

Murakami, M., et al., 2021b. Living with COVID-19: Mass gatherings and minimizing risk. QJM-Int. J. Med. 114, 437–439.

Nippon Professional Baseball, 2021. NPB 2021 season summary report: Season review under the influence of the COVID-19. https://npb.jp/npb/summaryreport2021_eng.pdf. (accessed 18 January, 2022).

Our World in Data, 2021. Coronavirus (COVID-19) vaccinations. https://ourworldindata.org/covid-vaccinations. (accessed 13 December 2021).

Parnell, D., et al., 2020. COVID-19, networks and sport. Manag. Sport Leis., doi: 10.1080/23750472.2020.1750100.

Prince-Guerra, J. L., et al., 2021. Evaluation of abbott BinaxNOW rapid antigen test for SARS-CoV-2 infection at two community-based testing sites — Pima County, Arizona, November 3–17, 2020. MMWR Morb. Mortal. Wkly. Rep. 70, 100–105.

Puranik, A., et al., 2021. Comparison of two highly-effective mRNA vaccines for COVID-19 during periods of Alpha and Delta variant prevalence (preprint). medRxiv. 2021.08.06.21261707.

Revollo, B., et al., 2021. Same-day SARS-CoV-2 antigen test screening in an indoor mass-gathering live music event: a randomised controlled trial. Lancet Infect. Dis. 21, 1365–1372.

Sleat, D., et al., 2021. Are vaccine passports and covid passes a valid alternative to lockdown? BMJ. 375, n2571.

Smith, J. A. E., et al., 2021. Public health impact of mass sporting and cultural events in a rising COVID-19 prevalence in England (preprint). https://khub.net/documents/135939561/338928724/Public+health+impact+of+mass+sporting+and+cultural+events+in+a+rising+COVID-19+prevalence+in+England.pdf/05204895-1576-1ee7-b41e-880d5d6b4f17. (accessed 9 December 2021).

Stange, M., et al., 2021. SARS-CoV-2 outbreak in a tri-national urban area is dominated by a B.1 lineage variant linked to a mass gathering event. PLoS Pathog. 17, e1009374.

Subcommittee on Novel Coronavirus Disease Control, 2021. 11th meeting materials. https://www.cas.go.jp/jp/seisaku/ful/taisakusuisin/bunkakai/dai11/gijisidai.pdf. (accessed 9 December 2021). (in Japanese)

The United Kingdom Government, 2021a. Information on the Events Research Programme. https://www.gov.uk/government/publications/guidance-about-the-events-research-programme-erp-paving-the-way-for-larger-audiences-to-attend-sport-theatre-and-gigs-safely-this-summer/guidance-on-the-events-research-programme. (accessed May 31, 2021).

The United Kingdom Government, 2021b. Science Note - Emerging findings from studies of indicators of SARS-CoV-2 transmission risk at the Events Research Programme: environment, crowd densities and attendee behaviour. https://www.gov.uk/government/publications/events-research-programme-phase-ii-and-iii-findings/science-note-emerging-findings-from-studies-of-indicators-of-sars-cov-2-transmission-risk-at-the-events-research-programme-environment-crowd-densi. (accessed 18 January, 2022).

Ueki, H., et al., 2020. Effectiveness of face masks in preventing airborne transmission of SARS-CoV-2. mSphere. 5, e00637–20.

UK Health Security Agency, 2021. SARS-CoV-2 variants of concern and variants under investigation in England. Technical briefing: Update on hospitalisation and vaccine effectiveness for Omicron VOC-21NOV-01 (B.1.1.529). https://assets.publishing.service.gov.uk/government/uploads/system/uploads/attachment_data/file/1044481/Technical-Briefing-31-Dec-2021-Omicron_severity_update.pdf. (accessed 18 January, 2022).

Watanabe, T., et al., 2010. Development of a dose-response model for SARS coronavirus. Risk Anal. 30, 1129–38.

Yasutaka, T., et al., 2021. Assessment of COVID-19 risk and prevention effectiveness among spectators of mass gathering events (preprint). medRxiv. 2021.07.05.21259882.

Zhang, X., Wang, J., 2021. Dose-response relation deduced for coronaviruses from coronavirus disease 2019, severe acute respiratory syndrome, and Middle East respiratory syndrome: Meta-analysis results and its application for infection risk assessment of aerosol transmission. Clin. Infect. Dis. 73, e241–e245.

